# Association of Visual Impairment with Risk for Future Parkinson’s Disease

**DOI:** 10.1101/2021.01.09.21249187

**Authors:** Zhuoting Zhu, Wenyi Hu, Huan Liao, Zachary Tan, Yifan Chen, Danli Shi, Xianwen Shang, Xueli Zhang, Yu Huang, Honghua Yu, Wei Wang, Mingguang He, Xiaohong Yang

## Abstract

**Background:** Although visual dysfunction is one of the most common non-motor symptoms among patients with Parkinson’s disease (PD), it is not known whether such dysfunction predates the onset of clinical PD.

**Objectives:** To examine the association of visual impairment (VI) with the future development of PD in the UK Biobank Study.

**Methods:** The UK Biobank Study is one of the largest prospective cohort studies of health, enrolling over 500,000 participants aged 40-69 years between 2006 and 2010 across the UK. VI was defined as a habitual distance visual acuity (VA) worse than 0.3 LogMAR in the better-seeing eye. Incident cases of PD were determined by self report data, hospital admission records or death records, whichever came first. Multivariable Cox proportional hazard regression models were used to investigate the association between VI and the risk of incident PD.

**Results:** A total of 117,050 participants were free of PD at the baseline assessment. During the median observation period of 5.96 (interquantile range [IQR]: 5.77-6.23) years, PD occurred in 222 (0.19%) participants. Visually impaired participants were at a higher risk of developing PD than non-VI participants (p<0.001). Compared with the non-VI group, the adjusted hazard ratio was 2.28 (95% CI 1.29-4.04, p=0.005) in the VI group. These results were consistent in the sensitivity analysis, where incident PD cases diagnosed within one year after the baseline assessment were excluded.

**Conclusions:** This prospective cohort study found that VI was associated with an increased risk of incident PD, suggesting that VI may represent a prodromal feature of PD.

## Introduction

Parkinson’s disease (PD) is the second most prevalent neurodegenerative disease and affects 2-3% of the global population aged over 65^1^. The total burden of disease associated with PD is increasing due to ageing populations, longer disease durations and environmental and social risk factors^2^. Relatively conservative estimates predict over 12 million patients will be affected by PD globally by 2050^3^.

In addition to motor symptoms, clinical features of PD involve many non-motor symptoms exacerbating overall disability^4^. Non-motor symptoms of PD have recently received increased attention as they precede the onset of motor symptoms, and may act as markers of preclinical stages of PD^5^. Visual dysfunction has been reported as one of the most common non-motor symptoms, and increasing evidence has demonstrated higher prevalence of this non-motor symptom in PD patients compared to healthy controls^6-9^. It is yet unknown however whether visual impairment (VI), an important indicator of visual dysfunction^10^, predates the onset of clinical PD. Further, understanding the association between VI and future risk of PD may facilitate early diagnosis and intervention of PD prior to manifestation of the full clinical syndrome.

In this context, we sought to examine the association of VI with risk of future PD development using the UK Biobank, a community-based longitudinal sample.

## Methods

### Study Sample

Detailed procedures of the UK Biobank study have been described elsewhere^11^. Briefly, more than 500,000 participants aged between 40-69 years were recruited at 22 assessment centers across the UK from 2006 to 2010. Data on lifestyle, environment and medical history were obtained using self-administered questionnaires. Physical measurements and biological specimens including blood, urine and saliva samples were also collected. Detailed follow-up of the health of participants was achieved through linkage to national electronic health record datasets. Ophthalmic examinations for baseline assessment were carried out from 2009 at six assessment centers. Participants with available visual acuity (VA) data (n=117,252) were included in the present analysis. Baseline characteristics of participants with and without VA data is shown in Supplement Table 1. Relative to participants without VA data, those with available VA data were more likely to be slightly older (56.8 vs. 56.4 years), of non-Caucasian ethnicity, less materially deprived, non-smokers, non-drinkers, physically active, diabetic, hypertensive, and less likely to use psychotropic medications. For analysis of incident PD, participants diagnosed with PD prior to baseline assessment (n=174), and those with self-reported PD at the baseline nurse interview (n=28) were excluded, leading to a final cohort with 117,050 participants included for analysis.

**Table 1.**
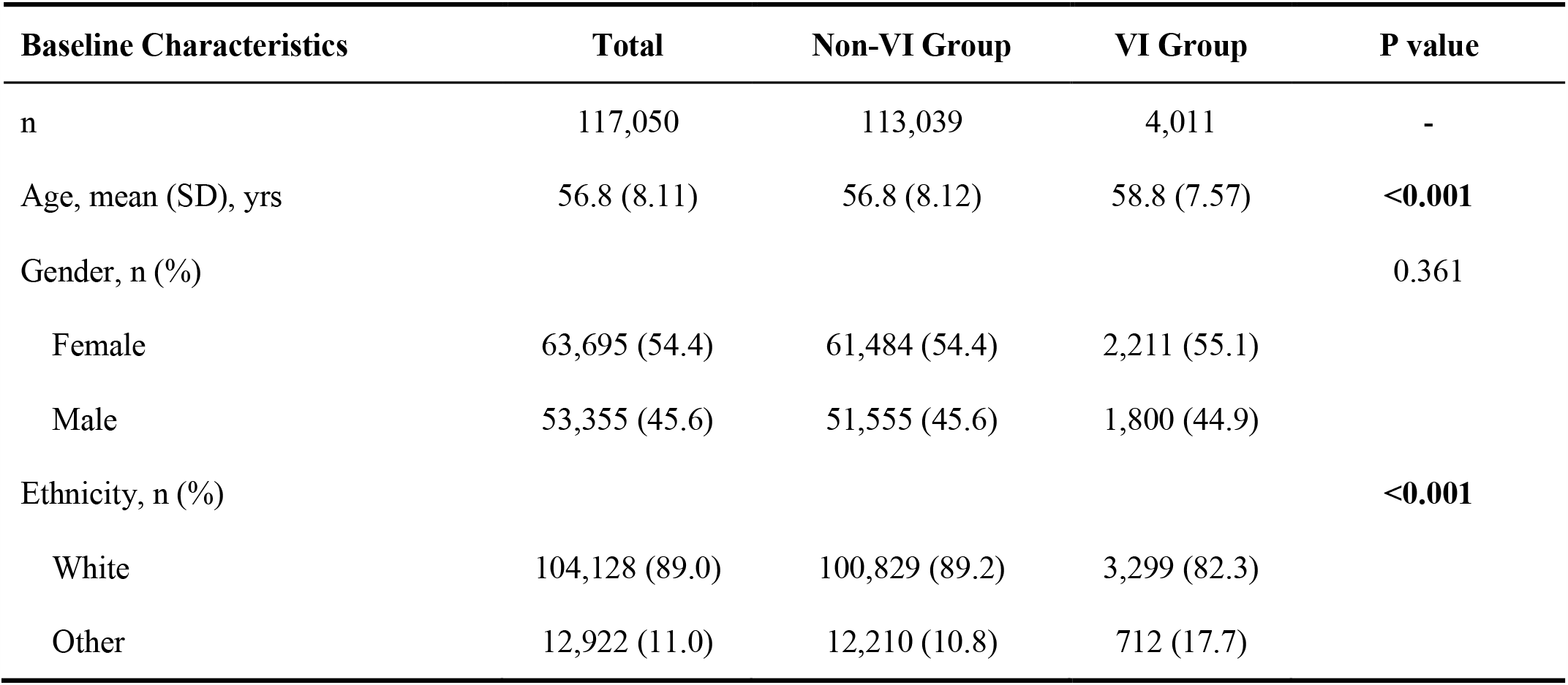
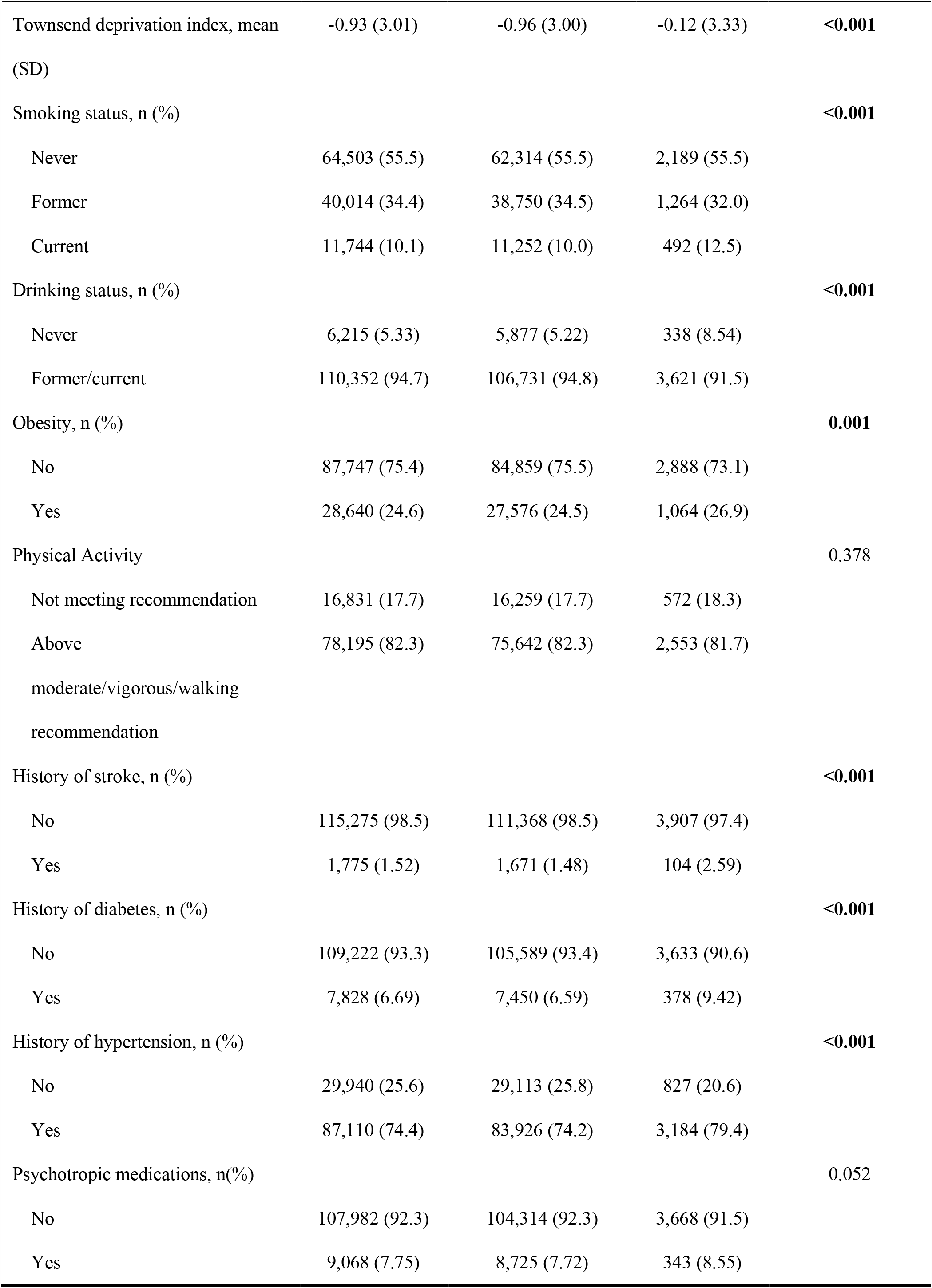
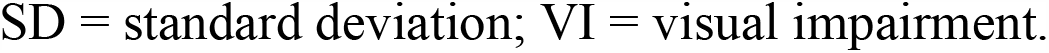
Baseline Characteristics of the Study Participants Stratified by Visual Impairment Status

### Standard Protocol Approvals, Registrations, and Patient Consents

Access to the UK Biobank data was granted after registration. The application ID was 62443. The UK Biobank has obtained Research Tissue Bank approval from its Research Ethics Committee recommended by the National Research Ethics Service (reference 11/NW/0382). Participants provided written informed consent.

### Assessment of Visual Acuity

Detailed methods for VA testing in the UK Biobank study have been described elsewhere^12^. Habitual distance VA was measured using a logarithm of the minimum angle of resolution (LogMAR) chart (Precision Vision, LaSalle, Illinois, USA). Participants were tested at 4 meters with habitual correction if any, or at 1 meter if participants were unable to read. Participants were required to read letters sequentially from the top and the total number of correctly identified letters was converted to a logMAR VA. Presenting VA worse than 0.3 logMAR units (equivalent to Snellen 20/40) was defined as VI.

### Ascertainment of PD

The algorithm for ascertaining PD in the UK Biobank Study could be found at in detail elsewhere^13^. In brief, ascertainment of PD was based on self-reported data, diagnosis in the hospital admissions data, or cause of death recorded in the national death register, whichever was the earliest. Hospital admission data were retrieved from Health Episode Statistics records (for participants from England and Wales) and the Scottish Morbidity Records (for participants from Scotland). The National Health Service (NHS) Information Centre, and the NHS Central Register Scotland were used to obtain the date and causes of death for participants from England and Wales, and Scotland, respectively. The UK Biobank study conducted a validation study on the accuracy of code sources for identification of PD cases, which achieved a positive predictive value (PPV) of 0.91 (95% confidence interval [CI], 0.83-0.96) when combining all sources (self report, hospital admission records, and death records)^13^. Follow-up length was calculated as the time interval between the date of baseline assessment and the censor date, which is either the date of PD diagnosis, death, lost to follow-up or 1^st^ March 2016.

### Covariates

Consistent with previous studies^14-17^, covariates for the analysis of incident PD included age at baseline assessment (continuous), sex (male/female), ethnicity (Caucasian and non-Caucasian), the Townsend deprivation index (continuous), smoking (current smoker, past smoker, and never), alcohol consumption (current and past/never drinker), obesity (yes/no), physical activity (above moderate/vigorous/walking recommendation, and not), history of stroke (yes/no), history of diabete mellitus (yes/no), history of hypertension (yes/no), and the use of psychotropic medications (yes/no).

Obesity was defined as body mass index (BMI) ≥30 kg/m^2^. History of diabete mellitus was defined to include participants with either doctor-diagnosed diabetes, insulin treatment or oral antidiabetic medications. History of hypertension was defined to include participants with either doctor-diagnosed hypertension, antihypertensive agents or measured systolic pressure ≥130 mmHg or diastolic pressure ≥80 mmHg. The previous use of anti-depressive, anti-migraine and axiolytic medications was defined as the use of psychotropic medications.

### Statistical Analysis

We reported normally distributed variables as means and standard deviation (SD); skewed variables as median and interquartile range (IQR), and categorical variables as number and percentages. Unpaired t-test and Pearson’s χ^2^ test were used to compare continuous and categorical variables, respectively. The log-rank test was used for comparison of incident PD distributions between VI and non-VI groups. The risks of incident PD associated with VI were evaluated using Cox proportional hazards models for estimation of hazard ratio (HR) with 95% confidence interval (CI).

To minimize the possibility of including prevalent cases in the present analysis, a sensitivity analysis was performed to exclude PD cases diagnosed in the year immediately following baseline assessment. Schoenfeld residuals were used to test the proportional hazards assumptions for Cox models and all variables were found to be valid. Two-sided p values less than 0.05 were considered statistically significant. All tests were conducted using Stata version 13 (StataCorp LLC, College Station, Texas USA).

### Data Availability Statement

The UK Biobank is an open-access resource to researchers through registration of proposed research. This present study was approved and registered as study 62443 with the UK Biobank resource.

## Results

A total of 117,050 participants (mean [SD] age: 56.8 [8.11] years; female: 63,695 [54.4%]) who were free of PD at the baseline assessment were included in this present analysis. Baseline characteristics of included participants are described in Table 1. A total of 4,011 (3.43%) participants suffered from VI. The visually impaired participants tended to be older, of non-Caucasian ethnicity, more materially deprived, current smokers, non-drinkers, obese, and with a history of stroke, diabetes and hypertension. There were no differences in other charateristics between the VI and non-VI group.

During the median follow up period of 5.96 (IQR: 5.77-6.23) years, a total of 222 cases of PD were recorded. The incidence of PD in all participants was 0.19%, with 16 (7.21%) being in the VI group and 206 (92.8%) being in the non-VI group, respectively. The log-rank test demonstrated a significant difference in the incidence of PD between the VI and non-VI groups (p<0.001). Baseline characteristics stratified by the incidence of PD are shown in Table 2. The incident PD group consisted of a greater proportion of participants that were of older age, male gender, non-smokers, less physically active, with a history of hypertension, and use of psychotropic medications at the baseline assessment compared to the non-PD group.

**Table 2.** Baseline Characteristics Stratified by Incident Parkinson Disease

| Baseline Characteristics | Non-PD Group | PD Group | P value |
| --- | --- | --- | --- |
| N | 116,828 | 222 | - |
| Age, mean (SD), yrs | 56.8 (8.11) | 63.2 (5.42) | **<0.001** |
| Gender, n (%) |  |  | **<0.001** |
| Female | 63,608 (54.5) | 87 (39.2) |  |
| Male | 53,220 (45.5) | 135 (60.8) |  |
| Ethnicity, n (%) |  |  | 0.452 |
| White | 103,927 (89.0) | 201 (90.5) |  |
| Others | 12,901 (11.0) | 21 (9.46) |  |
| Townsend deprivation index, mean (SD) | -0.93 (3.01) | -1.16 (2.89) | 0.258 |
| Smoking status, n (%) |  |  | **0.010** |
| Never | 64,389 (55.5) | 114 (51.4) |  |
| Former | 39,919 (34.4) | 95 (42.8) |  |
| Current | 11,731 (10.1) | 13 (5.86) |  |
| Drinking status, n (%) |  |  | 0.123 |
| Never | 6,198 (5.33) | 17 (7.66) |  |
| Former/current | 110,147 (94.7) | 205 (92.3) |  |
| Obesity, n (%) |  |  | 0.153 |
| No | 87,591 (75.4) | 156 (71.2) |  |
| Yes | 28,577 (24.6) | 63 (28.8) |  |
| Physical Activity |  |  | **0.007** |
| Not meeting recommendation | 16,786 (17.7) | 45 (25.4) |  |
| Above moderate/vigorous/walking recommendation | 78,063 (82.3) | 132 (74.6) |  |
| History of stroke, n (%) |  |  | 0.369 |
| No | 115,058 (98.5) | 217 (97.8) |  |
| Yes | 1,770 (1.52) | 5 (2.25) |  |
| History of diabetes, n (%) |  |  | 0.054 |
| No | 109,022 (93.3) | 200 (90.1) |  |
| Yes | 7,806 (6.68) | 22 (9.91) |  |
| History of hypertension, n (%) |  |  | **0.023** |
| No | 29,898 (25.6) | 42 (18.9) |  |
| Yes | 86,930 (74.4) | 180 (81.1) |  |
| Psychotropic medications, n(%) |  |  | **<0.001** |
| No | 107,798 (92.3) | 184 (82.9) |  |
| Yes | 9,030 (7.73) | 38 (17.1) |  |

SD = standard deviation; PD = Parkinson's disease.

The risks of developing PD for participants with and without VI based on Cox proportional hazard regression models are shown in Table 3. After adjusting for age, gender and ethinicity, the presence of VI was significantly associated with a higher risk of developing PD (HR=2.13, 95% CI: 1.27-3.56, p=0.004). The multivariable Cox proportional hazard regression model indicated that VI was associated with a 2.28-fold higher risk of developing subsequent PD (95% CI: 1.29-4.04, p=0.005). Similar findings were observed in the sensitivity analyses which excluded participants with PD cases diagnosed in the year immediately following baseline assessment (HR=1.93, 95% CI: 1.07-3.49, p=0.029, Table 3).

**Table 3.** Cox Proportional Hazards Models for Incident Parkinson Disease by Visual Impairment Status

| VI Status | Age-, Gender- and Ethnicity-Adjusted |  | Multivariable Model <sup>a</sup> |  |
| --- | --- | --- | --- | --- |
|  | Model |  |  |  |
|  | HR (95% CI) | P-value | HR (95% CI) | P-value |
| **Main analysis** |  |  |  |  |
| VI status |  |  |  |  |
| None | 1 [Reference] | - | 1 [Reference] | - |
| VI | **2.13 (1.27-3.56)** | **0.004** | **2.28 (1.29-4.04)** | **0.005** |
| **Sensitivity analysis <sup>b</sup>** |  |  |  |  |
| VI status |  |  |  |  |
| None | 1 [Reference] | - | 1 [Reference] | - |
| VI | **1.79 (1.05-3.03)** | **0.031** | **1.93 (1.07-3.49)** | **0.029** |

VI = visual impairment; PD = Parkinson's disease; HR = hazard ratio; CI = confidence interval.
<sup>a</sup> Cox proportional hazards regression models adjusted for age, gender, race/ethnicity, Townsend
deprivation index, smoking status, drinking status, obesity, physical activity, history of stroke, history
of diabetes, history of hypertension, and psychotropic medications.
<sup>b</sup> Excluding incident PD diagnosed in the year immediately following baseline assessment.

## Discussion

In this large community-based sample, we found that visually impaired individuals had a 2.28 times greater risk of developing PD compared to those who did not have any VI. Our findings suggest that VI may represent a prodromal feature of PD.

To the best of our knowledge, this is the first study to demonstrate that visually impaired individuals are at a greater risk of developing future PD than those without. Of note, it is well established that visual dysfunctions are common in PD. In a cross-sectional study of the European population, PD patients had a greater than double increase in the odds of having fair or poor eyesight^9^. A case-control study suggested that patients with mild to moderate PD had poorer performance compared to control subjects in visual perception tasks^6^. Worsened visual function as measured by visual acuity was also identified in PD patients in a cross-sectional study^7^.

Even though the mechanisms underlying the association of VI and incident PD were unclear, several plausible explanations had been raised. Firstly, the ‘bottom-up theory’ postulated that impairment of the retinal dopaminergic system might explain visual abnormalities in PD^18^. Both animal and clinical studies had suggested that dopaminergic retinal cell loss was commonly seen in PD^19-22^. Further, aggregates of α-synuclein in the retina that mimiced the pathological process of PD, had been reported to cause neurodegeneration of the dopaminergic cells in the retina^23^. Secondly, the ‘top-down’ theory might explain visual dysfunctions observed in PD. Support for this hypothesis came from evidence which demonstrated specific dysfunctions of the visual cortex in PD^24^. The depletion of neurotransmitters such as GABA and dopamine, as well as altered lipid metabolism in the visual cortex of PD patients, could account for visual symptoms^25-27^. Thirdly, strong evidence in associations between ocular diseases and PD might also account for the association between VI and the development of future PD^28^. Previous studies found that ocular diseases, such as glaucoma, cataract, age-related macular degeneration and diabetic retinopathy^29-31^, shared similar pathogenesis^32, 33^, risk factors^18, 34^ and proteomics profiles^35^ with PD. Further studies are warranted to confirm these speculations.

Our findings have an important implication for research and public health policy. In the present study, VI was identified as a potential prodromal feature of PD and might serve as a novel marker for the preclinical stages of PD, potentially enabling improved early diagnosis and management for those at higher risk of subsequently developing the disease. VA screening may be expanded beyond its current scope in the prevention of sight-threatening disease.

The strengths of the present study include its large sample size, long-term follow-up duration, and comprehensive adjustments for confounding factors. However, several limitations should be considered. Firstly, the present analysis was solely based on baseline VA. Further studies are needed to examine the impact of vision loss and/or other aspects of visual dysfunction on future risk of PD. Secondly, we did not have sufficient information to differentiate between causes of VI, thus preventing us from investigating associations between specific causes of VI and the risk of developing PD. Thirdly, due to the nature of an observational study, the causal relationship between VI and incident PD cannot be confirmed. Thirdly, the UK Biobank participants were relatively younger and might not represent the whole population. Nevertheless, this would not affect the association between VI and incident PD^36^. Lastly, we could not completely exclude the possibility of residual confounding.

## Conclusions

In summary, we found that VI was associated with a significantly increased risk of developing PD. Our findings highlighted the importance of vision screening in identifying individuals at high risk of developing PD. Further studies are needed to confirm our results, and the causal nature of VI and PD.

## Abbreviations and Acronyms

PD: Parkinson’s disease
VI: visual impairment
VA: visual acuity
LogMAR: logarithm of the minimum angle of resolution
NHS: National Health Service
IQR: interquartile range
HR: hazard ratios
CI: confidence intervals
SD: standard deviations
PPV: positive predictive value
BMI: body mass index

## Acknowledgements

The present work was supported by the Fundamental Research Funds of the State Key Laboratory of Ophthalmology, Project of Investigation on Health Status of Employees in Financial Industry in Guangzhou, China (Z012014075), Science and Technology Program of Guangzhou, China (202002020049). Prof. Mingguang He receives support from the University of Melbourne at Research Accelerator Program and the CERA Foundation. The Centre for Eye Research Australia receives Operational Infrastructure Support from the Victorian State Government. The sponsor or funding organization had no role in the design or conduct of this research.

## Appendix 1. Author Contributions

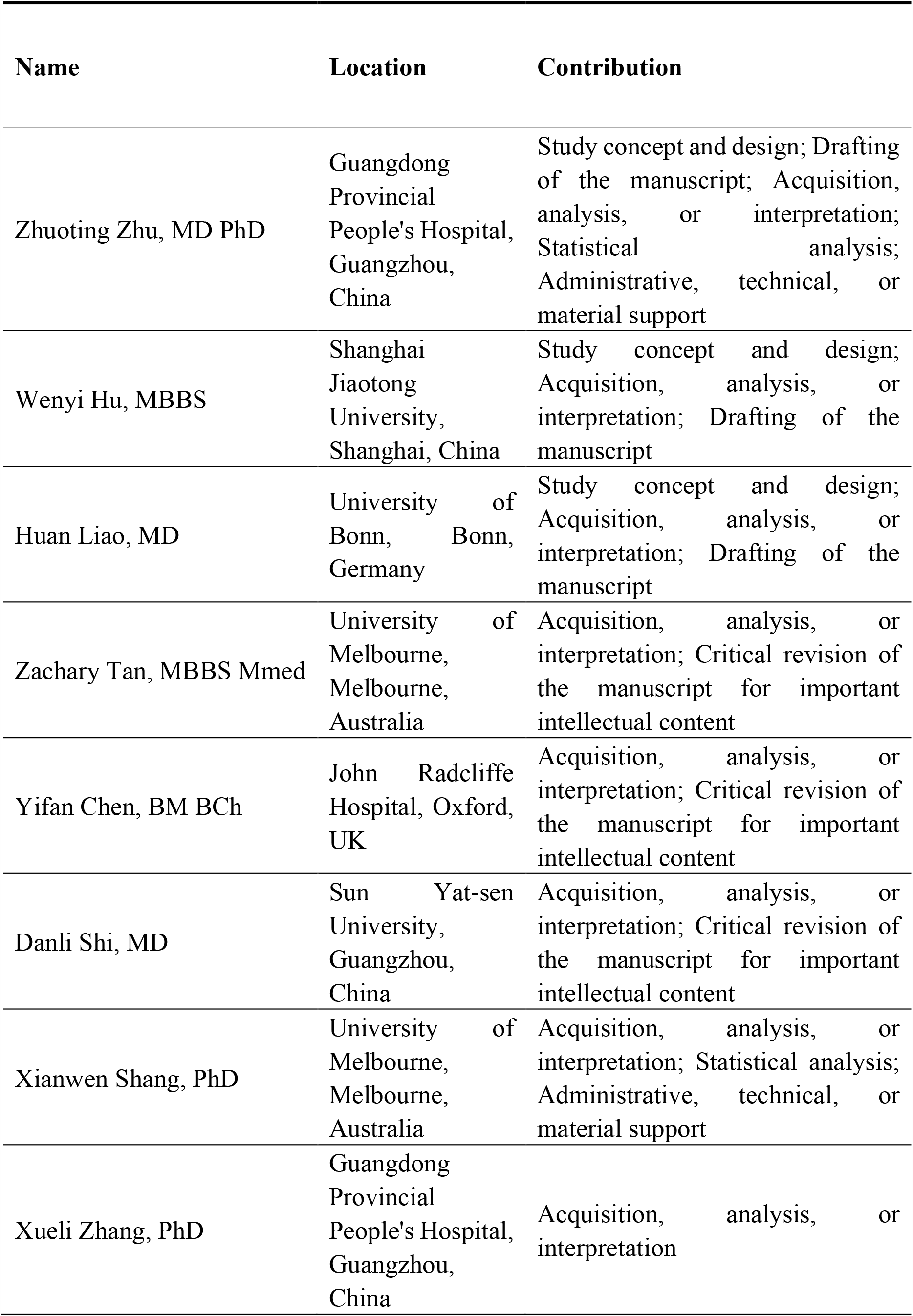
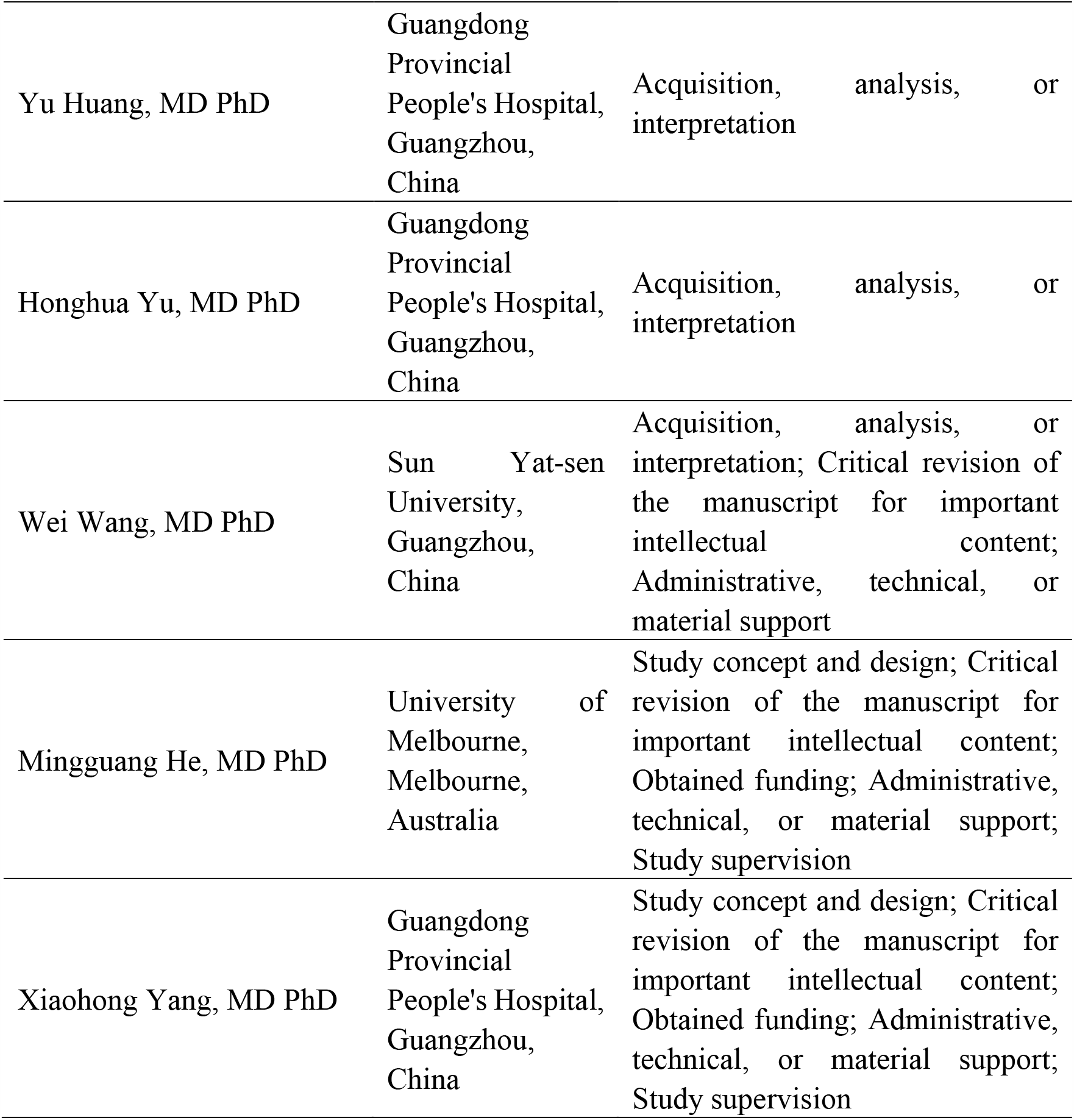

