## Supplement Table 1 for "Association of Visual Impairment with Risk for Future Parkinson’s Disease"

Supplementary Table 1. Baseline Characteristics of Participants With and Without Visual Acuity Data in the UK Biobank Study

| **Baseline Characteristics** | **No. of Participants without VA Data** | **No. of Participants with VA Data** | **P Value** |
|---|---|---|---|
| N | 385,252 | 117,252 | - |
| Age, mean (SD), yrs | 56.4 (8.09) | 56.8 (8.11) | **<0.001** |
| Gender, No. (%) | | | 0.890 |
| Female | 209,613 (54.4) | 63,769 (54.4) | |
| Male | 175,639 (45.6) | 53,483 (45.6) | |
| Ethnicity, No. (%) | | | **<0.001** |
| White | 368,385 (95.6) | 104,310 (89.0) | |
| Others | 16,868 (4.38) | 12,942 (11.0) | |
| Townsend index, mean (SD), yrs | -1.40 (3.11) | -0.93 (3.01) | **<0.001** |
| Smoking status, No. (%) | | | **<0.001** |
| Never | 208,893 (54.5) | 64,629 (55.5) | |
| Former | 132,981 (34.7) | 40,075 (34.4) | |
| Current | 41,221 (10.8) | 11,757 (10.1) | |
| Drinking status, n (%) | | | **<0.001** |
| Never | 16,163 (4.21) | 6,222 (5.33) | |
| Former/current | 367,920 (95.8) | 110,546 (94.7) | |
| Obesity, n (%) | | | 0.269 |
| No | 289,247 (75.6) | 87,905 (75.4) | |
| Yes | 93,566 (24.4) | 28,681 (24.6) | |
| Physical Activity | | | **<0.001** |
| Not meeting recommendation | 57,718 (18.8) | 16,874 (17.7) | |
| Above moderate/vigorous/walking recommendation | 249,368 (81.2) | 78308 (82.3) | |
| History of stroke, n (%) | | | 0.412 |

| | | | |
|---|---|---|---|
| No | 379,258 (98.4) | 115,467 (98.5) | |
| Yes | 5,995 (1.56) | 1,785 (1.52) | |
| History of diabetes, n (%) | | | **<0.001** |
| No | 362,834 (94.2) | 109,409 (93.3) | |
| Yes | 22,419 (5.82) | 7,843 (6.69) | |
| History of hypertension, n (%) | | | **<0.001** |
| No | 109,205 (28.4) | 29,984 (25.6) | |
| Yes | 276,048 (71.7) | 87,268 (74.4) | |
| Psychotropic medications, n(%) | | | **<0.001** |
| No | 353,379 (91.7) | 108,145 (92.2) | |
| Yes | 31,874 (8.27) | 9,107 (7.8) | |

SD = standard deviation
